# Predicting Active Surveillance Failure for Prostate Cancer Patients in the MRI Era: a Multicentre Transatlantic Cohort Study

**DOI:** 10.1101/2025.04.23.25326291

**Authors:** Nikita Sushentsev, Irene G. Li, George Xu, Anne Y. Warren, Celeste Y. Hsu, Madison Baxter, Dev Panchal, Christof Kastner, Sean Fernando, Ekaterina Pazukhina, Oleg Blyuss, Alexey Zaikin, Ahmed Shabaik, Anders M. Dale, Michael Liss, Tristan Barrett, Tyler M. Seibert

## Abstract

**Background and Objective:** MRI-driven active surveillance (AS) is increasingly used for prostate cancer (PCa) management. To determine the oncological safety of contemporary, MRI-driven AS and identify patients at higher risk of AS failure.

**Methods:** This retrospective cohort study included AS patients with MRI-localised PCa from three US and UK centres. The primary outcome was AS failure, a composite of PCa-specific mortality, metastasis, progression to ≥GG4, or post-treatment biochemical recurrence. The secondary outcome was disease progression, defined as histological progression to GG3 or progression to locally advanced disease. Hazard ratios (HRs) were estimated using multivariable Cox models, with multiplicity-adjusted log-rank tests comparing event-free survival across subgroups.

**Key Findings and Limitations:** 719 patients (median follow-up 5.2 years) were included. Of those, 629 (87%) had stable disease; 36 (5%) experienced AS failure, including 8 (1%) cases of metastasis and no PCa-related deaths; 54 (8%) had disease progression. Cribriform GG2 histology was the strongest predictor of AS failure (HR 12.7 95% CI, 4.8–33.6; *P* < 0.001), followed by tumor MRI-visibility (HR 5.0; 95% CI, 1.5-16.5; *P* = 0.009) and non-cribriform GG2 histology (HR 3.4; 95% CI, 1.6-7.0; *P* = 0.001). MRI-invisible, non-cribriform GG2 and all GG1 tumors had comparable event-free survival (*P*_*adj*_ > 0.05 for both outcomes). The study is limited by retrospective design.

**Conclusions and clinical implications:** Contemporary MRI-driven AS is safe, including for patients with non-cribriform GG2 tumours, particularly those that are MRI-invisible. Conversely, patients with cribriform GG2 disease are at increased risk of AS failure and therefore warrant upfront treatment.

## 1 Introduction

Prostate cancer (PCa) is among the most common and deadliest malignancies in Europe and US [1]. As minimising overtreatment is critical to early detection programmes, active surveillance (AS) is increasingly adopted to avoid or defer treatment-related side-effects while maintaining a curative intent [2].

While some guidelines limit AS to patients with low-risk PCa [3], some extend eligibility to intermediate-favourable risk patients with grade group (GG) 2 disease [4]. Hence, a higher proportion of patients now commence AS with MRI-visible, radiologically localised, GG2 disease [5] – a significant demographic shift from earlier low-risk AS cohorts with excellent outcomes [6].

To evaluate whether AS outcomes remain favourable in this evolving population, and whether there are patient subgroups requiring closer monitoring or upfront treatment, we conducted a retrospective cohort study across three academic centres in the UK and US.

## 2 Patients and methods

### 2.1 Study design and cohort characteristics

This retrospective cohort study was approved by the institutional review boards of the Cambridge University Hospitals (CUH/18/3592), University of California San Diego (191878), and University of Texas San Antonio (HSC20150160H). We included men enrolled on AS between March 2012 and March 2024 and meeting pre-defined inclusion criteria (**Figure 1, A**). Exclusion criteria were: <12 months of follow-up, participation in a therapeutic clinical trial during AS, or any missing baseline data. Follow-up included serial prostate-specific antigen (PSA) monitoring, MRI scans, and protocol-driven or triggered repeat biopsies. This study followed the Strengthening the Reporting of Observational Studies in Epidemiology (<u>STROBE</u>) guideline.

**Figure 1.**
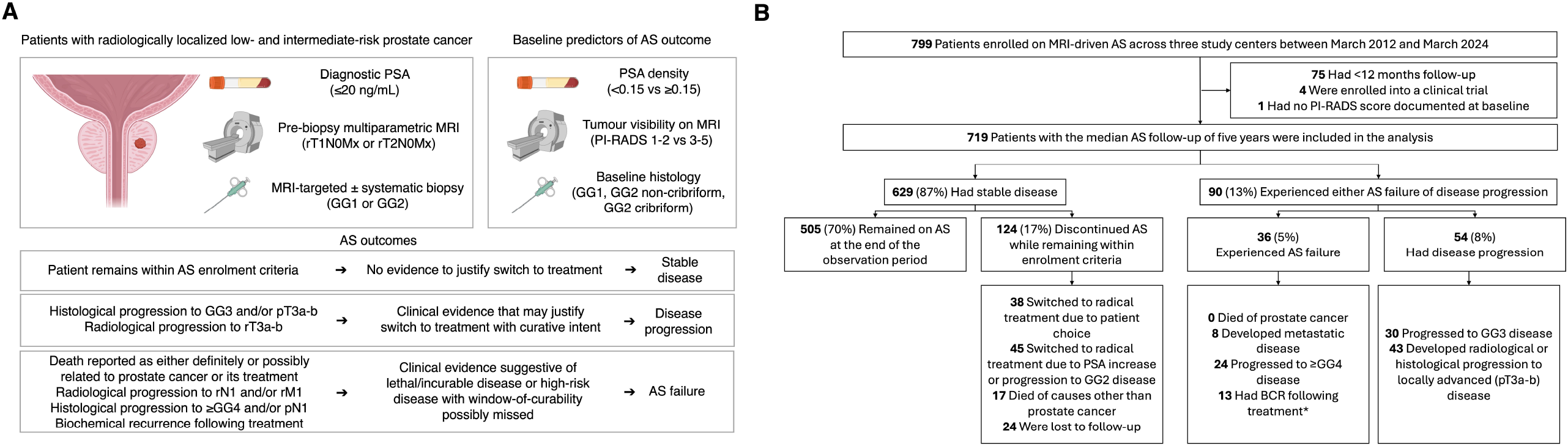
Study Overview and CONSORT Diagram. **(A)** Summary of the study inclusion criteria, exposures, and outcomes. **(B)** CONSORT diagram. AS indicates active surveillance; GG, grade group; MRI, magnetic resonance imaging; PI-RADS, Prostate Imaging-Reporting and Data System; PSA, prostate-specific antigen.

### 2.2 Outcomes and Exposures

The primary outcome was AS failure, a composite of oncological outcomes that indicate AS lost its intended benefit of deferring treatment while maintaining expectation of cure with initial definitive therapy. These were: death from PCa, radiological progression to metastatic disease, histological progression to GG≥4 disease, or biochemical recurrence following treatment (Phoenix definition [7]) (**Fig. 1A**). A secondary outcome, disease progression, included histological progression to GG3 disease or histological/definitive radiological progression to locally advanced disease. The disease progression endpoint reflects disease evolution that, in most cases, would justify commencing radical treatment with curative intent (**Fig. 1A**).

Key exposures included baseline biochemical (PSA density), radiological (MRI-visibility: PI-RADS scores 3-5 *vs*. PI-RADS scores 1-2), and histological (biopsy GG and cribriform status) predictors of AS outcomes commonly featured in major clinical guidelines [3,9,10]. Cribriform status was defined as the presence of any amount of invasive cribriform carcinoma and/or intraductal carcinoma, according to the relevant International Society of Urological Pathology [ISUP] guidelines [8].

### 2.3 Statistical analysis

Multivariable Cox proportional hazards (PH) models estimated hazard ratios (HRs) and 95% confidence intervals (CIs) for AS failure and disease progression. Right censoring was applied at the last AS follow-up date for stable patients or those discontinuing AS for non-progression reasons. For metastasis or biochemical recurrence occurring after definitive treatment, AS failure was recorded as occurring at date of initial treatment, which may have altered the natural history of disease but failed to cure it. For patients to be considered recurrence-free, they had to undergo at least 36 months of post-treatment follow-up. False discovery rate-adjusted (Hold-Šidak; *α* = 0.05) log-rank tests compared Kaplan-Meier survival curves across subgroups. The analysis was conducted in MATLAB (version 2023b) and GraphPad Prism (version 10.4.1), with a 2-sided P < 0.05 considered statistically significant.

## 3 Results

### 3.1 Baseline characteristics and AS outcomes

By March 2024, 799 patients were enrolled on MRI-driven AS across the three study centers (**Fig. 1B**). After excluding 80 patients, 719 remained. Of those, 36/719 (5%) men experienced AS failure, including 8/719 (1%) cases of metastasis and no PCa-related deaths; a further 54/719 (8%) men had disease progression, with the detailed cohort characteristics shown in **Table 1**. Of 629/719 (87%) patients with stable disease followed up for a median of 5.2 years (IQR 3.1-7.8 years), 124 men discontinued AS despite meeting the enrolment criteria (**Fig. 1B)**, thereby bringing the final AS retention rate to 505/719 (70%).

**Table 1.**
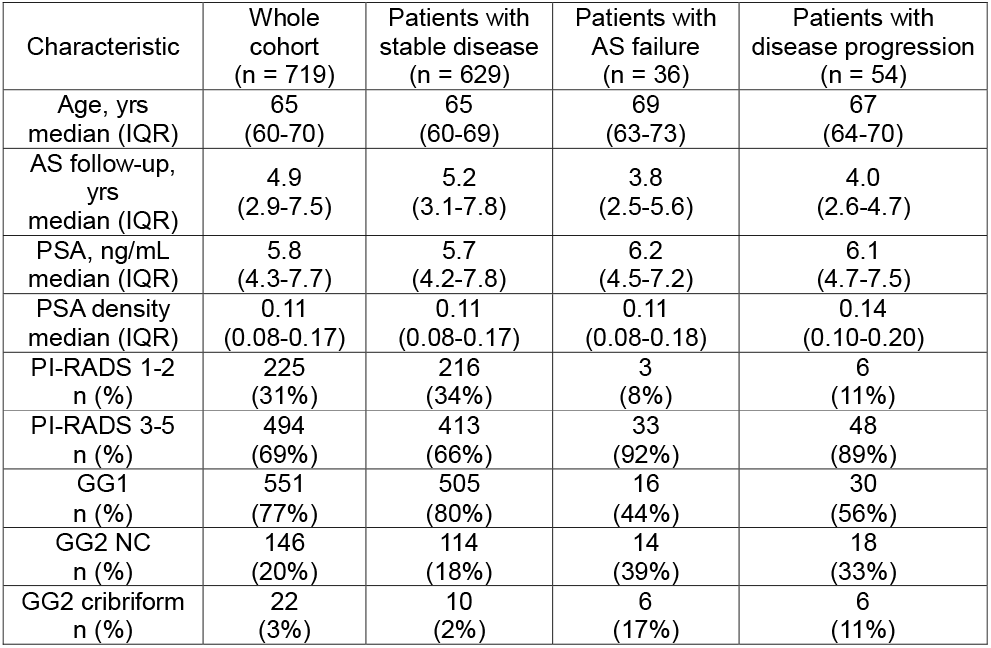
Baseline Cohort Characteristics. AS indicates active surveillance, GG grade group, IQR interquartile range, NC non-cribriform, PI-RADS Prostate Imaging-Reporting and Data System score, PSA prostate-specific antigen. Abbreviations: AS, active surveillance, GG, grade group, IQR, interquartile range, NC, non-cribriform, PI-RADS, Prostate Imaging-Reporting and Data System, PSA, prostate-specific antigen.

### 3.2 Baseline predictors of AS failure

The HR for AS failure was 12.7 times greater (95% CI, 4.8-33.6; *P* < 0.001) for patients with cribriform GG2 tumours than for those with GG1 tumours, compared to the 3.4-fold increased risk (95% CI, 1.6-7.0; *P* = 0.001) for non-cribriform GG2 disease (**Fig. 2A**). Patients with MRI-visible tumours had a 5.0-fold HR increase for AS failure (95% CI, 1.5-16.5; *P* = 0.009), whereas increased PSA density had no significant effect (HR, 1.02; 95%CI, 0.5-2.1; *P* = 0.947).

**Figure 2.**
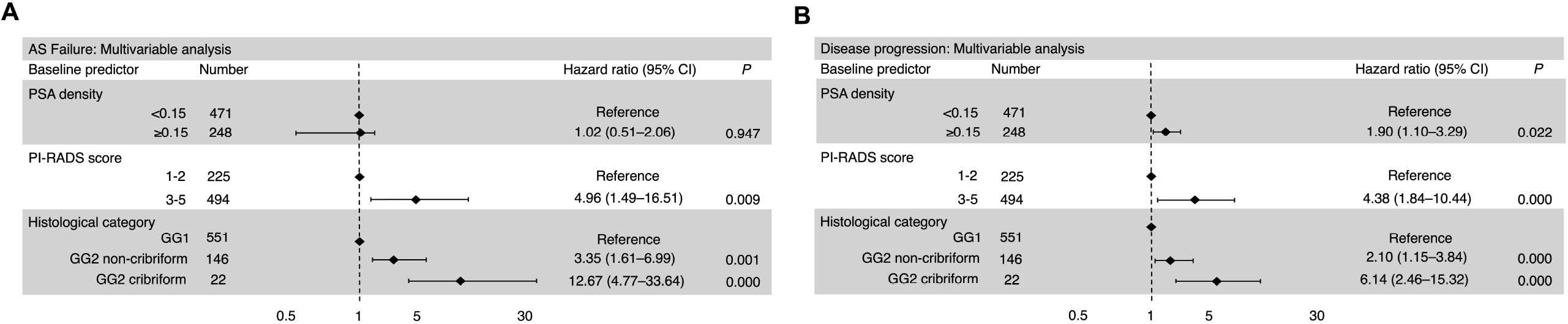
Outcome Analysis. Data are from multivariable Cox proportional hazards regression for AS failure **(A)** and disease progression **(B)**, diamonds indicate hazard ratios, and horizontal lines indicate 95% CIs. AS indicates active surveillance; GG, grade group; MRI, magnetic resonance imaging; PI-RADS, Prostate Imaging-Reporting and Data System; PSA, prostate-specific antigen.

Subgroup survival analysis revealed that cribriform GG2 lesions—all of which were MRI-visible—yielded the shortest AS failure-free survival compared to all other groups (*P* < 0.05 for all; **Fig. 3A**). Patients with MRI-visible, non-cribriform GG2 disease had the second-highest risk, with significantly shorter failure-free survival compared to all GG1 patients (*P*_adj_ < 0.05 for all; **Fig. 3A**). Notably, all patients with GG1 and MRI-invisible, non-cribriform GG2 disease had comparable failure-free survival (*P*_adj_ > 0.05 for all; **Fig. 3A**).

**Figure 3.**
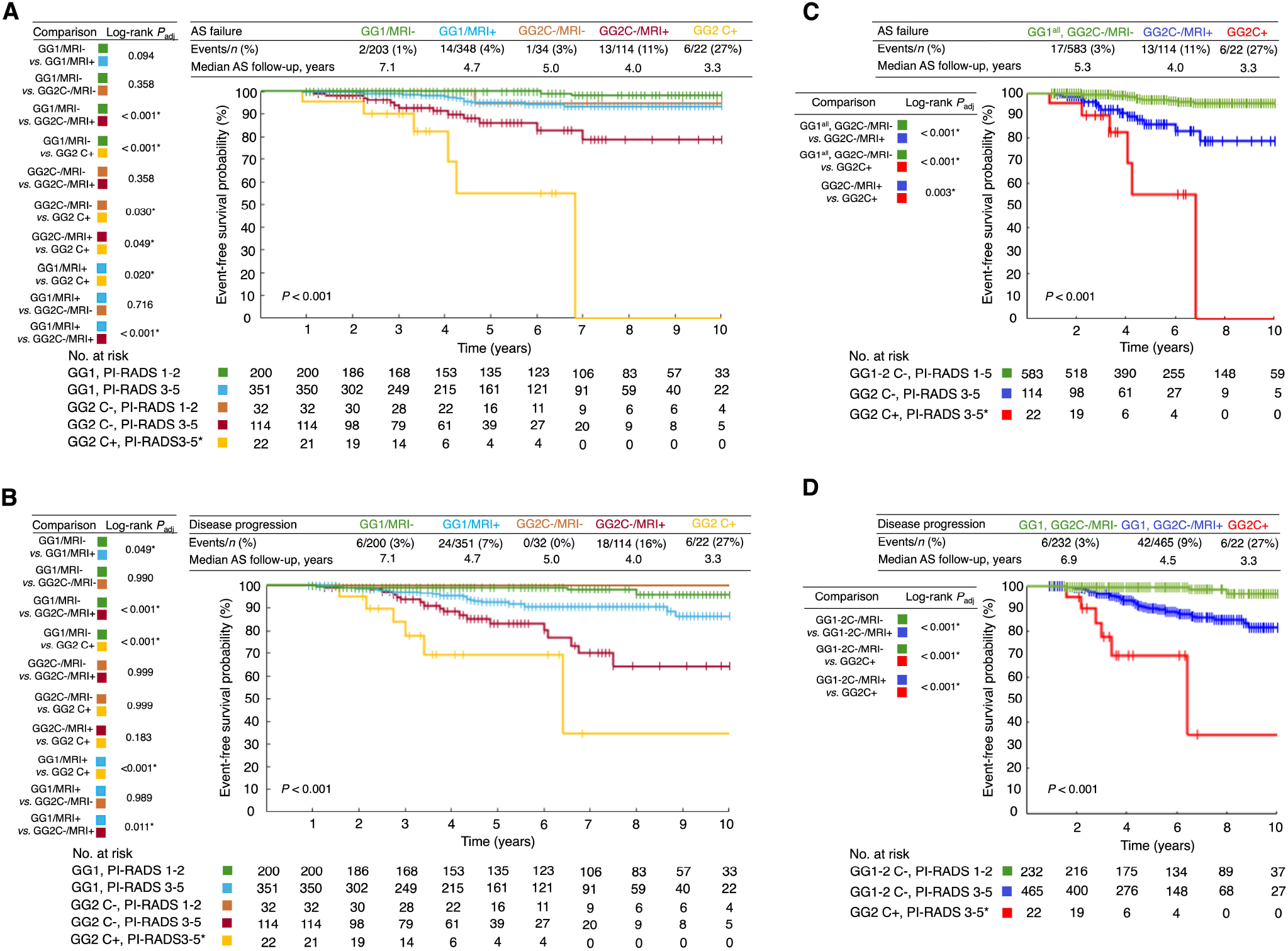
Subgroup Analysis of Outcomes Among Patients with Different Histological and Radiological Tumor Characteristics. Data are from a secondary, false discovery rate-adjusted Kaplan-Meier survival analysis. In panel (C), GG1^all^, GG2C-/MRI-includes all patients with GG1 disease and patients with MRI-invisible, non-cribriform GG2 disease; GG2C-/MRI+ indicates patients with MRI-visible, non-cribriform GG2 disease; GG2 C+ indicates patients with GG2 cribriform disease. In panel (**D)**, GG1, GG2C-/MRI-indicates patients with MRI-invisible GG1 and non-cribriform GG2 disease; GG1, GG2C-/MRI+ indicates patients with MRI-visible GG1 and non-cribriform GG2; GG2C+ indicates patients with GG2 cribriform disease. AS indicates active surveillance; C+, cribriform-positive; C-, cribriform-negative; GG, grade group; MRI, magnetic resonance imaging; MRI+, MRI-visible; MRI-, MRI-invisible; NC, non-cribriform; PI-RADS, Prostate Imaging-Reporting and Data System; PSA, prostate-specific antigen *All patients with GG2 cribriform disease also had MRI-visible (PI-RADS 3-5) disease.

### 3.3 Baseline predictors of disease progression

Similar overall trends were observed for disease progression, where increased PSA density also showed a significant association (HR, 1.9; 95% CI, 1.1-3.3; *P* = 0.02; **Fig. 2B**). Moreover, patients with MRI-visible GG1 had significantly shorter disease progression-free survival compared to GG1 patients with MRI-invisible disease (*P*_adj_ < 0.05; **Fig. 3B**).

### 3.4. Risk groups for AS failure and disease progression

Consequently, three distinct risk subgroups were identified for AS failure: cribriform GG2; MRI-visible, non-cribriform GG2; and all others, including MRI-invisible GG2 and all GG1 (*P*_adj_ < 0.05 for all; **Fig. 3C**). For disease progression, the three groups were: cribriform GG2; MRI-visible, non-cribriform GG1-2; MRI-invisible, non-cribriform GG1-2 (*P*_adj_ < 0.05 for all; **Fig. 3D**).

## Discussion

Overall, at an AS failure-rate of 5% (including 1% rate of metastases and no PCa-related deaths), our findings indicate excellent medium-term AS outcomes for patients with low- and intermediate-favourable-risk disease who were all diagnosed, staged, and followed-up using MRI. As expected, patients with GG1 disease at baseline were most likely to remain on AS. Those with MRI-invisible, non-cribriform GG2 disease had outcomes indistinguishable from those with GG1 disease. Conversely, patients with MRI-visible, non-cribriform GG2 disease (who would now constitute the majority of AS cohorts in centres that omit biopsy in patients with negative MRI [5]) had higher risk of AS failure, perhaps warranting closer surveillance. Dynamic risk-adjusted predictive models are currently being developed for this group [11].

While MRI-visibility and GG2 have been previously identified as important predictors of AS progression [12–15], the definition of progression has varied significantly across the studies. Many studies of AS use GG upgrade as an endpoint, but this is questionable when upgrade from GG1 to GG2 does not exclude the patient from continued AS eligibility—and may not even portend a worse prognosis at all, given the results presented here. While death from PCa and progression to metastasis are undoubtedly the strongest and most meaningful endpoints, there are other outcomes increasingly used in recent studies to encompass AS failure. These include post-treatment disease recurrence (representing a failure to deliver on the intention to cure the disease with initial therapy) and progression to high-risk (GG≥4) disease which would involve significant treatment escalation compared to intermediate-risk disease. By combining these four outcomes into one composite endpoint, we show that 11% of patients with MRI-visible, non-cribriform disease experience some form of AS failure (after median 5.2 years of follow-up), highlighting the need for additional biomarkers that would enable early detection of progression to guide treatment of these men.

In contrast, we show that cribriform GG2 morphology is the strongest predictor of adverse AS outcomes, with 27% of such patients experiencing AS failure. Although the current European guidelines already exclude patients with cribriform disease from AS, these recommendations are largely based on historic, radically treated cohorts [10], as opposed to contemporary cohorts managed with AS. In addition, these recommendations are not reflected in the North American or UK guidelines which, together with substantial variation in AS practices, means that many patients with cribriform disease may be currently offered AS. To our knowledge, our findings provide the first direct evidence of adverse AS outcomes in patients with cribriform disease and support recommending upfront treatment to these men.

Considering the significant variation of AS practices and common inclusion of GG2 patients in AS programmes, our definition of disease progression was limited to histological progression to GG3 disease or progression to locally advanced disease. By doing so, we mostly aimed at reducing bias associated with more subjective endpoints such as progression to treatment, where the decision is often driven by patients or clinicians in the absence of hard clinical evidence of evolving disease phenotype. Similar to AS failure, patients with cribriform GG2 disease detected at baseline were at the highest risk of disease progression, most of which occurred within the first four years of AS, indicating the lack of benefit of AS for these men.

The study was limited by its retrospective design and the inevitable lack of standardisation of AS protocols across the three centres representing different healthcare systems. Future prospective studies identifying appropriate AS schedules and novel predictors of AS failure for patients with MRI-visible, non-cribriform GG2 will help maximise the safety and efficiency of contemporary AS programmes. That said, considering the existing European guidelines and the growing recognition of the adverse nature of cribriform morphology strengthened by this study, prospective studies further investigating AS outcomes in patients with cribriform disease are unlikely to be supported.

In conclusion, this study demonstrates the safety of MRI-driven AS at a median follow-up of five years, while providing direct evidence supporting upfront treatment of patients with cribriform GG2 disease. Our results also suggest that low-intensity surveillance is likely adequate for not only GG1 disease but also non-cribriform GG2 disease if the tumor is not MRI-visible.

## Data Availability

All data produced in the present study are available upon reasonable request to the authors

## Funding Acknowledgments

The study was funded by the National Institute for Health and Care Research (NIHR), UK, and Cancer Research UK Cambridge Centre. The views expressed are those of the authors and not necessarily those of the NIHR or the Department of Health and Social Care. N.S. acknowledges support from Emmanuel College, Cambridge. A.Y.W. is supported by the Urological Malignancies Programme of the Cancer Research UK Cambridge Centre (C9685/A25177) and NIHR Cambridge Biomedical Research Centre (BRC-1215-20014). TMS acknowledges support from the Prostate Cancer Foundation (24CHAL03).

